# Chronotropic Incompetence Limits Aerobic Exercise Capacity in Patients Taking Beta-Blockers

**DOI:** 10.1101/2020.07.15.20149005

**Authors:** Maciej Tysarowski, Krzysztof Smarz, Beata Zaborska, Ewa Pilichowska-Paszkiet, Malgorzata Sikora-Frac, Andrzej Budaj, Tomasz Jaxa-Chamiec

## Abstract

**Background:** Chronotropic incompetence in patients taking beta-blockers is associated with poor prognosis; however, its impact on exercise capacity (EC) remains unclear.

**Methods:** We retrospectively analyzed data from consecutive patients taking beta-blockers referred for cardiopulmonary exercise testing. EC was expressed as peak oxygen uptake (peak VO2; mL/kg/min). Chronotropic incompetence was defined as chronotropic index (CI) ≤ 62%. CI was calculated as [(HR at peak–HR at rest) / (maximum predicted HR–HR at rest)] × 100%.

**Results:** Among 140 patients all taking beta-blockers (age 61 ± 9.7 years; 73% males), there were 113 (80.7%) patients with chronotropic incompetence. EC was lower in the group with chronotropic incompetence than the group without it, peak VO2 18.3 ± 5.7 vs. 24.0 ± 5.3 mL/kg/min, p < 0.001. In multivariate analysis EC correlated positively with CI (β = 0.14, p < 0.001) and male gender (β = 5.12, p < 0.001), and negatively with age (β = −0.17, p < 0.001) and presence of heart failure (β = −3.35, p < 0.001). Beta-blocker dose was not associated with EC. Partial correlation attributable to CI accounted for more than one-third of the variance in EC explained by the model.

**Conclusions:** In patients taking beta-blockers, presence of chronotropic incompetence was associated with lower EC, regardless of the beta-blocker dose. CI accounted for more than one-third of EC variance explained by our model.

**HIGHLIGHTS:**

- Patients on beta blockers with chronotropic incompetence have lower exercise capacity
- Beta-blocker dose is not associated with exercise capacity
- We recommend incorporating chronotropic index into exercise stress testing

## 1 INTRUDUCTION

Chronotropic incompetence is defined as an inadequate increase in heart rate (HR) during exercise. The presence of chronotropic incompetence is associated with poor prognosis in both symptomatic and asymptomatic patients.^1–6^ The frequency of chronotropic incompetence depends on the definition used as well as the population examined and ranges from 9% to 89%.^7^ The Euro-EX prevention trial reported that, among healthy individuals, chronotropic incompetence assessed as an inability to achieve 80% of the age-predicted HR reserve, was present in 70% of patients.^8^ While chronotropic incompetence in patients not treated with beta-blockers is correlated with reduced exercise capacity (EC),^9^ data from patients taking beta-blockers are ambiguous. Furthermore, in studies aimed at assessing chronotropic response and EC, chronotropic incompetence was most commonly defined as an inability to reach 80% of CI or 80% of the maximum predicted HR regardless of beta-blockers treatment, and only few of them were focused on patients treated with beta-blockers.^10–13^

Identifying factors responsible for low EC may be relevant for patient evaluation and management,^14 15^ because its strong predictive value for all-cause and cardiovascular mortality in patients with and also without heart failure.^16^ In a study of 3736 consecutive patients with normal electrocardiograms who were taking beta-blockers (either metoprolol or atenolol), chronotropic incompetence defined as a CI £ 62% was an independent predictor of all-cause mortality (adjusted hazard ratio 1.94, 95% confidence interval 1.43 to 2.64, p <0.0001) in 4.5 years follow-up.^3^ Based on this assumption, this cut-off value is recommended in the guidelines for chronotropic incompetence diagnosis in patients treated with beta-blockers,^16^ but this value has not been validated as a predictor of low EC.

Therefore, we planned our study to assess the relationship between chronotropic incompetence and EC in patients treated with beta-blockers.

## 2 METHODS

### 2.1 Study population and inclusion criteria

We retrospectively analyzed data from consecutive patients who were referred for an exercise tolerance assessment at the Exercise Physiology Laboratory at the Cardiology Department, Centre of Postgraduate Medical Education, Grochowski Hospital, Warsaw, Poland, between January 2008 and June 2016.

We included patients treated with beta-blockers starting at least 4 weeks before cardiopulmonary exercise testing as presented in Fig. 1. Exclusion criteria were pulmonary or peripheral limitations of exercise, hemodynamically significant valve dysfunction or pulmonary hypertension, heart failure in New York Heart Association functional class IV, hospitalization for acute coronary syndromes or decompensated heart failure within the past 30 days, exercise-induced ischemia, insufficient effort (respiratory exchange ratio < 1.05 at peak exercise), heart rhythm other than sinus during cardiopulmonary exercise test, and pacemaker implantation. Patients included in the study were divided into two groups: with and without chronotropic incompetence.

**Fig. 1.**
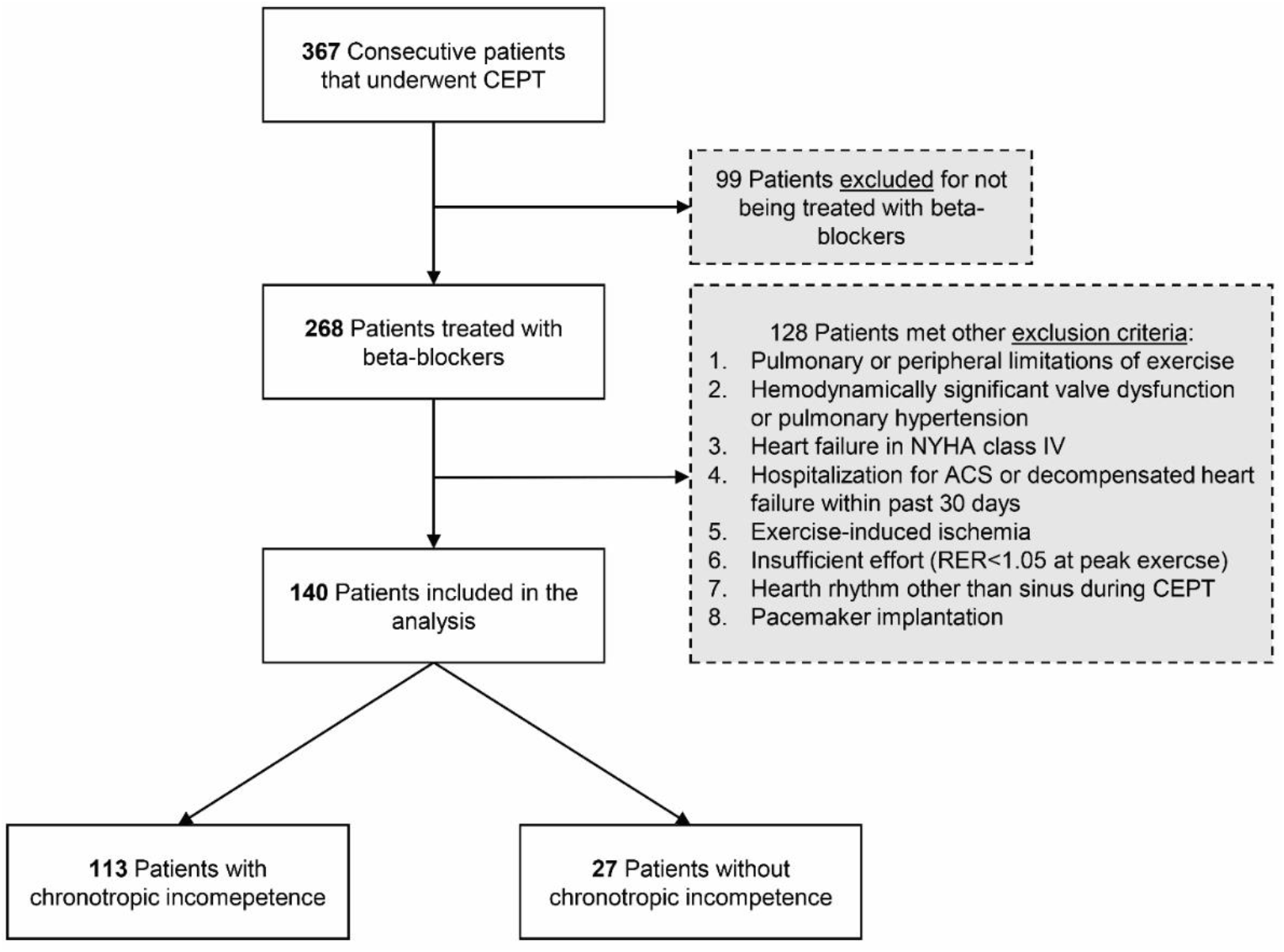
Study flow chart. Abbreviations: ACS, acute coronary syndrome; CPET, cardiopulmonary exercise test; NYHA, New York Heart Association functional classification; RER, respiratory exchange ratio.

Data on patients’ demographic and clinical details, laboratory tests, medications, and comorbidities were obtained from hospital patient medical documentation. Comorbidities were established based on physicians’ diagnoses from electronic medical records. Creatinine clearance was calculated using Cockcroft-Gault equation. Daily doses of beta-blockers were calculated as bisoprolol equivalent dose. Dose equivalents for beta-blockers were derived from the European Society of Cardiology Guidelines for the diagnosis and treatment of acute and chronic heart failure.^17^ Doses were calculated as bisoprolol 10 mg daily equivalent to: carvedilol 25 mg twice daily (BID); metoprolol tartrate 100 mg BID; metoprolol succinate 200 mg daily; nebivolol 10 mg daily; and sotalol 160 mg BID.

### 2.2 Echocardiography

All assessed echocardiographic studies were performed during routine evaluation by cardiologists experienced in cardiovascular imaging, and all measurements were performed according to recommendations of the American Society of Echocardiography and the European Association of Cardiovascular Imaging.^18–20^ Patients were characterized using the following echocardiographic parameters: left ventricular end-diastolic dimension; anteroposterior dimension of the left atrium from parasternal long-axis view; basal right ventricular end-diastolic dimension and minor-axis dimension of the right atrium from four-chamber view; left ventricular ejection fraction from the biplane method of discs (modified Simpson’s rule); visual assessment of segmental contraction disturbances as (1) normal or hyperkinetic, (2) hypokinetic, (3) akinetic, and (4) dyskinetic; calculated wall motion score index (16 segments model); left ventricular diastolic dysfunction with grade 1, 2, or 3 dysfunction based on mitral inflow parameters; and right ventricular systolic dysfunction diagnosed in patients with tricuspid plane systolic excursion <17 mm.

### 2.3 Cardiopulmonary exercise test

All patients performed a symptom-limited cardiopulmonary exercise test using a treadmill or cycle ergometer with Schiller Cardiovit CS-200 (Schiller, Baar, Switzerland) and Ergo Spiro adapter (Garnshorn, Nederlauer, Germany), with the incremental protocol selected according to the individual’s physical condition to maintain the duration of exercise between 8 and 12 min. All patients were familiar with the exercise protocol and were encouraged to perform maximal effort (≥8 points using the 10-point Borg scale). All exercise tests were performed and analyzed by the same physician according to the guidelines of the American College of Cardiology/American Heart Association and the American Thoracic Society/American College of Chest Physicians.^14 16 21 22^ The system was calibrated each day before performing the tests. Ventilation, VO_2_ uptake, and carbon dioxide output during exercise were analyzed breath by breath. Peak VO_2_ (mL/kg/min) was averaged from measurements taken during the last 20 s of exercise and was assessed as an EC parameter. The anaerobic threshold was calculated using a dual method approach (V-slope and ventilatory equivalent methods). Maximum predicted VO_2_ values were calculated using the Wasserman/Hansen equation.^23^

Chronotropic incompetence was defined as chronotropic index (CI) ≤ 62%,^3^ calculated as percentage of HR reserve as follows:

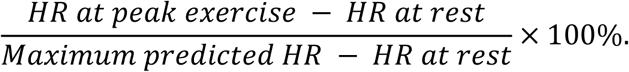

Maximum predicted HR was calculated as 220 − age, as previously defined by Astrand et al.^24^ Percent of predicted maximum HR achieved at peak exercise and HR reserve, defined as the change in HR from rest to peak were also calculated.

Other analyzed cardiopulmonary exercise testing parameters included systolic blood pressure at rest and at peak exercise, ventilatory efficiency (minute ventilation versus carbon dioxide production slope), and breathing reserve at peak exercise, calculated as the percentage of maximal voluntary ventilation used [(maximal voluntary ventilation − minute ventilation at peak exercise)/maximal voluntary ventilation] × 100%. Resting spirometry parameters, calculated as forced expiratory volume in 1 s., and inspiratory vital capacity were also recorded.

### 2.4 Statistical methods

Data were presented as mean ± standard deviation for normally distributed or median (25th and 75th percentiles) for non-normally distributed continuous variables. Categorical variables were presented as a number (percentage). Normality for all continuous variables was tested using the Shapiro–Wilk test. Group comparisons were performed using Student’s *t* test or Mann–Whitney test for continuous variables and χ^2^ (chi-squared) test for categorical variables. Univariate and multivariate linear regression analyses were performed to establish the association between independent variables and EC. Peak VO_2_ (mL/kg/min) was used as the dependent variable for all models. Logarithmic transformation was used for non-normally distributed variables when analyzed in regression models. Variables for the univariate and multivariate models were selected using the Akaike Information Criterion and stepwise linear regression model. Variables with well-known effects on EC were forced into the model. All statistical tests were two-sided. Statistical significance was established as α = 0.05, and all statistical analyses were performed using R Statistical Software version 3.6.1.

### 2.5 Ethics

This study was conducted following the requirements set out in the Declaration of Helsinki. All patients provided written consent to take part in the cardiopulmonary exercise test. The study protocol was approved by the Center of Postgraduate Medical Education’s Institutional Review Board, and individual consent to participate in retrospective anonymous data analysis was waived.

## 3 RESULTS

Among 367 evaluated patients, 268 were treated with beta-blockers and 140 fulfilled the inclusion criteria. The flow chart of patient selection is presented in Fig. 1.

The baseline characteristics of the patients are presented in Table 1. Among 140 patients all taking beta-blockers, mean age was 61 ± 9.7 years and 73% were males. There were 113 (81%) patients in the group with chronotropic incompetence and 27 (19%) patients in the group without chronotropic incompetence. Diabetes mellitus and impaired fasting glucose were more frequent in the group with chronotropic incompetence. Daily dose of beta-blockers was higher, and serum creatinine levels were lower in the group with chronotropic incompetence, and there were no differences in creatinine clearance between the two groups. No statistically significant differences were observed for other demographic and clinical parameters between the two groups.

**Table 1.** Baseline characteristics of study participants.

|  | All patients<br>(n = 140) | Chronotropic incompetence<sup>a</sup> |  | p-value |
| --- | --- | --- | --- | --- |
|  |  | Yes<br>(n = 113) | No<br>(n = 27) |  |
| Demographics |  |  |  |  |
| Age, years | 61.0 ± 9.7 | 60.7 ± 10.0 | 62.1 ± 8.5 | 0.525 |
| Male sex, n (%) | 102 (73) | 78 (69) | 24 (89) | 0.065 |
| BMI, kg/m<sup>2</sup> | 27.8 ± 4.1 | 27.8 ± 4.0 | 27.7 ± 4.5 | 0.953 |
| Comorbidity, n (%) |  |  |  |  |
| NYHA functional class | 89 (64) | 74 (65) | 15 (56) | 0.514 |
| I | 71 (51) | 59 (52) | 12 (44) | 0.609 |
| II | 8 (6) | 7 (6) | 1 (4) | 0.968 |
| III | 10 (7) | 8 (7) | 2 (7) | 0.968 |
| CAD | 115 (82) | 94 (83) | 21 (78) | 0.704 |
| MI | 110 (79) | 90 (80) | 20 (74) | 0.709 |
| Coronary angiography | 118 (84) | 96 (85) | 22 (81) | 0.88 |
| PCI | 107 (76) | 87 (77) | 20 (74) | 0.945 |
| CABG | 5 (4) | 4 (4) | 1 (4) | 1 |
| DM/IFG | 29 (20) | 28 (24) | 1 (4) | **0.033** |
| Hypertension | 88 (63) | 74 (65) | 14 (52) | 0.273 |
| Current smoker | 42 (30) | 35 (31) | 7 (26) | 0.779 |
| Paroxysmal atrial fibrillation | 14 (10) | 12 (11) | 2 (7) | 0.886 |
| Biochemistry |  |  |  |  |
| Hemoglobin, g/L | 13.8 ± 1.5 | 13.8 ± 1.5 | 13.8 ± 1.5 | 0.934 |
| Serum creatinine (IQR), mg/dL | 0.9 (0.8–1.1) | 0.9 (0.8–1.1) | 1.1 (0.9–1.3) | **0.005** |
| Creatinine clearance, mL/min/1.73 m<sup>2</sup> | 95 ± 30 | 97 ± 31 | 88 ± 29 | 0.188 |
| **Medication, n (%)** |  |  |  |  |
| Bisoprolol | 77 (55) | 67 (59) | 10 (37) | 0.061 |
| Metoprolol | 49 (35) | 35 (31) | 14 (52) | 0.069 |
| Carvedilol | 9 (6) | 7 (6) | 2 (7) | 1 |
| Sotalol | 2 (1) | 2 (2) | 0 (0) | 1 |
| Nebivolol | 2 (1) | 1 (1) | 1 (4) | 0.837 |
| Other heart rate lowering drugs<sup>b</sup> | 8 (6) | 16 (14) | 2 (7) | 1 |
| Dihydropyridine CCB | 18 (13) | 16 (14) | 2 (7) | 0.554 |
| ACEI/ARB | 111 (79) | 92 (81) | 19 (70) | 0.313 |
| Diuretics | 35 (25) | 30 (27) | 5 (19) | 0.536 |
| BB dose, bisoprolol equivalent (IQR), mg | 2.5 (2.5–5) | 2.5 (2.5–5.0) | 2.5 (1.2–2.5) | **0.033** |
| **Echocardiography** |  |  |  |  |
| LV end-diastolic dimension, mm | 47 (44–50) | 46.0 (44–50) | 47 (44–50) | 0.523 |
| Left atrium dimension, mm | 40 ± 5 | 40 ± 5 | 39 ± 5 | 0.558 |
| LVEF, % | 53.3 ± 11.6 | 53.4 ± 11.9 | 52.8 ± 10.5 | 0.816 |
| WMSI (IQR) | 1.4 (1.1–1.7) | 1.4 (1.1–1.8) | 1.5 (1.0–1.6) | 0.940 |
| LV diastolic dysfunction, n (%) |  |  |  | 0.702 |
| Grade 1 | 114 (81) | 90 (80) | 24 (89) | 0.404 |
| Grade 2 | 8 (6) | 7 (6) | 1 (4) | 0.968 |
| Grade 3 | 2 (1) | 2 (2) | 0 (0) | 1 |
| MR moderate, n (%) | 33 (24) | 25 (22) | 8 (30) | 0.567 |
| RV end-diastolic dimension (IQR), mm | 34 (30–36) | 34 (29–36) | 35 (32–38) | 0.096 |
| Right atrium dimension, mm | 35 ± 6 | 36 ± 6 | 34 ± 5 | 0.267 |
| RV systolic dysfunction, n (%) | 38 (27) | 33 (29) | 5 (19) | 0.378 |
| TR moderate, n (%) | 11 (8) | 9 (8) | 2 (7) | 1 |

Note: Values represent mean ± SD, median (IQR; 25<sup>th</sup> – 75<sup>th</sup> percentiles), or number (%). Bold values indicate statistical significance.
Abbreviations: ACEI / ARB, angiotensin-converting enzyme inhibitor/angiotensin receptor blocker; BB, beta-blocker; BMI, body mass index; CABG, coronary artery bypass grafting; CAD, coronary artery disease; CCB, calcium channel blocker; DM / IFG, diabetes mellitus / impaired fasting glucose; IQR, interquartile range; LV, left ventricle; LVEF, left ventricular ejection fraction; MI, history of myocardial infarction; MR, mitral regurgitation; NYHA, *New York Heart Association* functional classification; PCI, percutaneous coronary intervention; RV, right ventricle; TR, tricuspid regurgitation; WMSI, wall motion score index.
<sup>a</sup>defined as chronotropic index ≤62%.
<sup>b</sup>non-dihydropyridine calcium channel blockers, amiodarone, propafenone, ivabradine, and digoxine

The echocardiographic parameters are also presented in Table 1. No differences were observed for left and right ventricular function and chambers dimensions between the two groups. Median time between echocardiography and cardiopulmonary exercise testing was 4 weeks.

Cardiopulmonary exercise testing results are shown in Table 2. The CI for all patients was 48.6% ± 17.3%, lower in the group with chronotropic incompetence 42.7% ± 13.0% vs. 73.1% ± 9.3% in the group without chronotropic incompetence. The group with chronotropic incompetence also had a significantly lower percentage of maximum predicted HR achieved at peak exercise compared with the group without chronotropic incompetence.

**Table 2.** Cardiopulmonary exercise testing parameters of study participants.

|  | All patients<br>(n = 140) | Chronotropic incompetence<sup>a</sup> |  | p-value |
| --- | --- | --- | --- | --- |
|  |  | Yes<br>(n = 113) | No<br>(n = 27) |  |
| Treadmill exercise test, n (%) | 109 (78) | 86 (76) | 23 (85) | 0.446 |
| Cycle ergometer exercise test, n (%) | 31 (22) | 27 (24) | 4 (15) | 0.446 |
| VO<sub>2</sub> at anaerobic threshold, L/min | 1.1 ± 0.4 | 1.1 ± 0.3 | 1.4 ± 0.4 | **<0.001** |
| VO<sub>2</sub> at anaerobic threshold, mL/kg/min | 13.7 ± 3.7 | 13.2 ± 3.7 | 16.0 ± 2.9 | **<0.001** |
| VO<sub>2</sub> at peak, L/min | 1.6 ± 0.6 | 1.5 ± 0.5 | 2.1 ± 0.7 | **<0.001** |
| VO<sub>2</sub> at peak, mL/kg/min | 19.4 ± 6.1 | 18.3 ± 5.7 | 24.0 ± 5.3 | **<0.001** |
| VO<sub>2</sub> at peak, mL/kg/min % predicted | 73 ± 19 | 69 ± 17 | 89 ± 18 | **<0.001** |
| CO<sub>2</sub> at peak, L/min | 1.8 ± 0.8 | 1.6 ± 0.7 | 2.4 ± 0.9 | **<0.001** |
| METs at peak | 5.5 ± 1.7 | 5.2 ± 1.6 | 6.9 ± 1.5 | **<0.001** |
| HR at rest (IQR), bpm | 72 (64–83) | 72 (64–82) | 77 (67–86) | 0.091 |
| HR at anaerobic threshold, bpm | 97 ± 13 | 95 ± 13 | 106 ± 10 | **<0.001** |
| HR at peak, bpm | 115 ± 17 | 110 ± 15 | 136 ± 10 | **<0.001** |
| HR at peak, % predicted | 72 ± 10 | 69 ± 8 | 86 ± 4 | **<0.001** |
| Chronotropic index, % | 48.6 ± 17.3 | 42.7 ± 13.0 | 73.1 ± 9.3 | **<0.001** |
| SBP at rest, mmHg | 127 ± 13 | 127 ± 13 | 130 ± 13 | 0.289 |
| SBP at peak (IQR), mmHg | 170 (155–180) | 170 (150–180) | 180 (160–190) | 0.075 |
| RER at peak (IQR) | 1.10 (1.05–1.16) | 1.09 (1.05–1.16) | 1.14 (1.06–1.21) | 0.072 |
| Min. ventilation vs CO<sub>2</sub> slope (IQR) | 24 (22–28) | 25 (23–28) | 23.6 (21–25) | 0.095 |
| Breathing reserve at peak (IQR), % | 45 (25–57) | 46 (44–50) | 47 (44–50) | 0.523 |
| FEV 1/IVC, % predicted | 93 ± 21 | 93 ± 20 | 94 ± 23 | 0.841 |

*Note:* Values represent mean ± SD, median (IQR; 25<sup>th</sup> – 75<sup>th</sup> percentiles), or number (%). Bold values indicate statistical significance.
*Abbreviations:* FEV 1/IVC, forced expiratory volume in the first second/inspiratory vital capacity; HR, heart rate; IQR, interquartile range; METs, metabolic equivalents; RER, respiratory exchange ratio; SBP, systolic blood pressure; VCO<sub>2</sub>, carbon dioxide production; VO<sub>2</sub>, oxygen uptake.
<sup>a</sup>defined as chronotropic index ≤62%.

EC expressed as peak VO_2_ was 19.4 ± 6.1 mL/kg/min for all patients, lower in the group with chronotropic incompetence compared with the group without chronotropic incompetence (18.3 ± 5.7 mL/kg/min vs. 24.0 ± 5.3 mL/kg/min, p < 0.001). The percentages of maximum predicted VO_2_ and VO_2_ at the anaerobic threshold were also significantly lower in the group with chronotropic incompetence. Peak VO_2_ was, 21.3 ± 5.3 mL/kg/min for males, and 14.3 ± 4.9 mL/kg/min for females, p < 0.001.

Fig. 2 shows there was a moderately strong (*r* = 0.55), positive linear association between CI and EC.

**Fig. 2.**
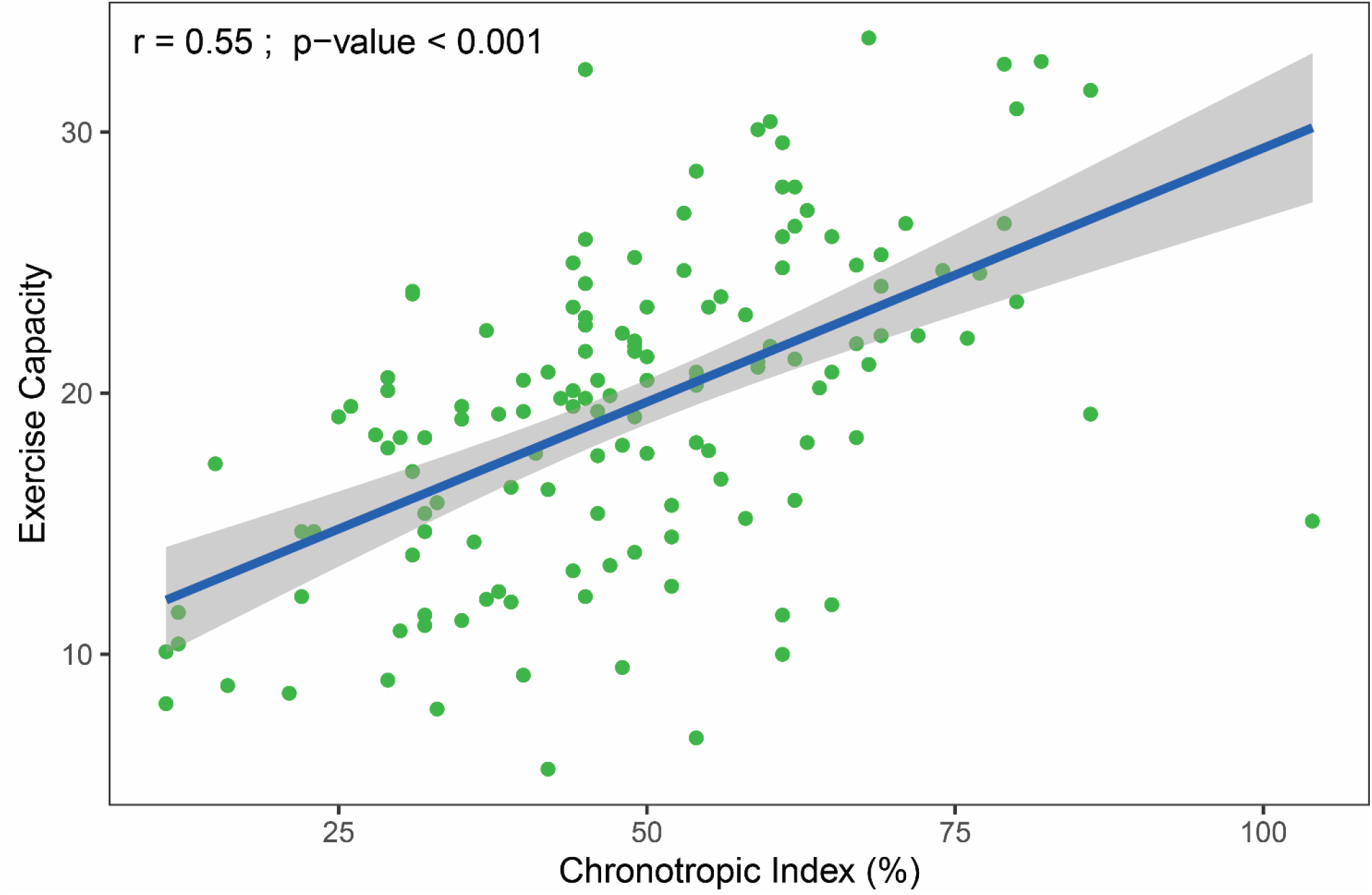
The relationship between exercise capacity (oxygen uptake at peak exercise in mL/min/kg) and chronotropic index. The gray-shaded area represents the standard error of regression line (blue). The r-value was calculated using Pearson correlation.

The regression analysis results are presented in Table 3. Univariate regression analysis revealed that CI, male gender, treadmill exercise testing, hemoglobin concentration, and peak systolic blood pressure correlated positively with EC, while age, presence of heart failure, and diabetes mellitus / impaired fasting glucose were negatively correlated.

**Table 3.** Results of univariate and multivariate linear regression analyses assessing predictors of exercise capacity (EC) measured as oxygen uptake at peak exercise (VO2peak, mL/min/kg).

|  | Univariate analysis |  |  | Multivariate analysis |  |  |  |
| --- | --- | --- | --- | --- | --- | --- | --- |
| | $\beta$ regression coefficient | 95% CI | p-value | $\beta$ regression coefficient | 95% CI | p-value | Explained variance (%)<sup>a</sup> |
| Chronotropic index, % | 0.20 | 0.15 to 0.24 | **<0.001** | 0.14 | 0.09 to 0.18 | **<0.001** | 24.7 |
| Male gender | 7.01 | 5.05 to 8.97 | **<0.001** | 5.12 | 2.86 to 7.38 | **<0.001** | 14.0 |
| Age, years | -0.22 | -0.32 to -0.13 | **<0.001** | -0.17 | -0.26 to -0.09 | **<0.001** | 12.9 |
| Heart failure | -4.55 | -6.52 to -2.60 | **<0.001** | -3.35 | -4.97 to -1.72 | **<0.001** | 11.8 |
| WMSI |  |  | 0.483 |  |  | 0.050 | 3.1 |
| Treadmill exercise test | 3.92 | 1.56 to 6.28 | **0.001** |  |  | 0.066 | 2.7 |
| BB daily dose, bisoprolol equivalent, mg |  |  | 0.743 |  |  | 0.140 | 1.8 |
| LVEF, % |  |  | 0.147 |  |  | 0.224 | 1.2 |
| Hemoglobin, g/L | 0.81 | 0.15 to 1.48 | **0.017** |  |  | 0.298 | 0.9 |
| Serum creatinine, mg/dL |  |  | 0.295 |  |  | 0.343 | 0.7 |
| LV diastolic dysfunction, grade 2 and 3 |  |  | 0.108 |  |  | 0.413 | 0.5 |
| Height, cm |  |  | 0.430 |  |  | 0.430 | 0.5 |
| DM / IFG | -3.25 | -5.70 to -0.80 | **0.010** |  |  | 0.560 | 0.3 |
| Current smoker |  |  | 0.670 |  |  | 0.586 | 0.2 |
| RV systolic dysfunction |  |  | 0.931 |  |  | 0.948 | 0.0 |
| SBP at peak exercise, mmHg | 0.05 | 0.00 to 0.09 | **0.034** |  |  |  |  |
| Hypertension |  |  | 0.137 |  |  |  |  |
| CAD |  |  | 0.133 |  |  |  |  |

Note: For multivariate analysis, R<sup>2</sup> = 64.1%, adjusted R<sup>2</sup> = 59.8%. Bold values indicate statistical significance.
Abbreviations: BB, beta-blocker; CAD, coronary artery disease; CI, confidence interval; DM/IFG, Diabetes mellitus/impaired fasting glucose; LV, left ventricle; LVEF, left ventricular ejection fraction; RV, right ventricle; SBP, systolic blood pressure; WMSI, wall motion score index.
<sup>a</sup>calculated as partial R<sup>2</sup>.

Multivariate analysis revealed positive correlations with EC which remained for CI and male gender, and negative correlations for age and presence of heart failure. Beta-blocker dose was not independently associated with EC. The partial correlation attributable to CI (partial R^2^ = 24.7%) accounted for more than one-third of the variance in EC explained by the model (model adjusted R^2^ = 59.8%)

## 4 DISCUSSION

This study revealed that in patients taking beta-blockers, CI was the strongest independent predictor of EC and accounted for one-third of the EC variability explained by the multivariable model. Other factors, such as gender, age, and presence of heart failure, were also independent predictors of EC, although to a lesser extent. Our model revealed that echocardiographic parameters, hemoglobin and creatinine levels, the modality of the exercise test, and beta-blocker dose had no discernible effects on EC among our study population. Our results confirmed previous findings that CI correlated linearly with peak VO_2_ (Fig. 1).^5^ To the best of our knowledge, this is the first study investigating which factors and to what extent can predict EC in patients taking beta-blockers.

Our findings also revealed significant differences in EC with regard to chronotropic incompetence defined as a CI cut-off value of 62%. This cut-off value has previously been shown to have prognostic significance in patients treated with beta-blockers.^3^ Age, gender, body mass index, physical activity, smoking and many comorbidities were previously examined as independently related to EC.^25^

Similarly to our study, the relationship between chronotropic incompetence and EC in 549 congestive heart failure patients taking beta-blockers was investigated by Magri et al. The authors concluded that chronotropic incompetence negatively correlates with EC regardless of beta-blocker daily dose. However in this study chronotropic incompetence was diagnosed as an inability to achieve 80% of CI or 80% of maximum predicted HR.^12^

Our results do not reveal correlation between beta-blocker dose and EC. Although it has been shown that high doses of beta-blockers can cause, aggravate, or reveal latent chronotropic incompetence,^7 26 27^ beta-blocker cessation did not normalize chronotropic response to exercise.^28^

Our study has some limitations as this was a single-center, observational, retrospective study with a relatively small group of mostly male patients. It included a diverse group of patients with and without heart failure. Brain natriuretic peptide plasma concentration values were not available in all patients and therefore were not included in the analyses. In the analyzed group, patients may have had chronotropic incompetence irrespective of, or induced by beta-blockers. It was also shown previously that endurance training during cardiac rehabilitation can improve chronotropic response in patients taking beta-blockers.^10^ Therefore, daily physical activity status can contribute in chronotropic response and EC. In our study data on physical activity status and quality of life were not available for all patients and were therefore not included in our analyses.

## 5 CONCLUSIONS AND CLINICAL IMPLICATIONS

In consecutive patients taking beta-blockers and referred for cardiopulmonary exercise testing, chronotropic incompetence, calculated as CI, was found to be an independent predictor of reduced EC. CI positively and independently correlated with EC with the highest explained variance. Therefore, we recommend that for patients taking beta-blockers, CI is incorporated as an exercise testing parameter, also in regular exercise stress tests. Our study revealed that beta-blocker dose was not an independent predictor of EC. Correlations between chronotropic response to exercise and beta-blocker daily doses need to be evaluated in further prospective studies. We are planning a long-term follow-up to evaluate predictors of mortality in the study group.

## Data Availability

The complete raw dataset can be accessed via the Mendeley Data repository below.

https://data.mendeley.com/datasets/w2jf6fr8hg/draft?a=4abca7bb-b565-4b03-bbee-f5598f56282d.

## ABBREVIATION LIST

CI: chronotropic index
EC: exercise capacity
HR: heart rate
VO_2_: oxygen uptake

## AUTHOR CONTRIBUTIONS

**Maciej Tysarowski**: conceptualization, methodology, software, formal analysis, resources, data curation, writing – review & editing, visualization. **Krzysztof Smarz**: conceptualization, methodology, investigation, validation, resources, data curation, writing – original draft, review & editing, visualization, project administration, funding acquisition. **Beata Zaborska**: investigation, resources, data curation. **Ewa Pilichowska-Paszkiet**: investigation, resources, data curation. **Malgorzata Sikora-Frac**: investigation, resources, data curation. **Andrzej Budaj**: writing – review & editing, supervision, project administration, funding acquisition. **Tomasz Jaxa-Chamiec:** writing – review & editing, supervision, project administration, funding acquisition.

All authors take responsibility for all aspects of the reliability and freedom from bias of the data presented and their discussed interpretation.

## DATA SHARING STATEMENT

The complete raw dataset can be accessed via the Mendeley Data repository: https://data.mendeley.com/datasets/w2jf6fr8hg/draft?a=4abca7bb-b565-4b03-bbee-f5598f56282d.

